# Capturing Global, Predicting Local for Controlling Antimicrobial Resistance: a retrospective multivariable analysis

**DOI:** 10.1101/2021.05.26.21257778

**Authors:** Raghav Awasthi, Vaidehi Rakholia, Samprati Agarwal, Lovedeep Singh Dhingra, Aditya Nagori, Tavpritesh Sethi

## Abstract

**Background:** Antimicrobial resistance (AMR) is a complex multifactorial outcome of health, socio-economic and geopolitical factors. Therefore, tailored solutions for mitigation strategies could be more effective in dealing with this challenge. Knowledge-synthesis and actionable models learned upon large datasets are critical in order to diffuse the risk of entering into a post-antimicrobial era.

**Objective:** This work is focused on learning *Global* determinants of AMR and predicting the susceptibility of antibiotics at the isolate level *(Local)* for WHO (world health organization) declared critically important pathogens Pseudomonas aeruginosa, Klebsiella pneumoniae, Escherichia coli, Acinetobacter baumannii, Enterobacter cloacae, Staphylococcus aureus.

**Methods:** In this study, we used longitudinal data (2004-2017) of AMR having 633820 isolates from 70 Middle and High-income countries. We integrated AMR data with the Global burden of disease (GBD), Governance (WGI), and Finance data sets in order to find the unbiased and actionable determinants of AMR. We chose a Bayesian Decision Network (BDN) approach within the causal modeling framework to quantify determinants of AMR. Finally Integrating Bayesian networks with classical machine learning approaches lead to effective modeling of the level of AMR.

**Results:** From MAR (Multiple Antibiotic Resistance) scores, we found that developing countries are at higher risk of AMR compared to developed countries, for all the critically important pathogens. Also, Principal Components Analysis(PCA) revealed that governance, finance, and disease burden variables have a strong association with AMR. We further quantified the impact of determinants in a probabilistic way and observed that health system access and government effectiveness are strong actionable factors in reducing AMR, which was in turn confirmed by what-if analysis. Finally, our supervised machine learning models have shown decent performance, with the highest on Staphylococcus aureus. For Staphylococcus aureus, our model predicted susceptibility to Ceftaroline and Oxacillin with the highest AUROC, 0.94(with SE of 0.01%) and 0.89(with SE of 0.002%) respectively.

## Introduction

Antimicrobial resistance is the reduction in the efficacy of antimicrobials in treating infections due to pathogen evolution under strong selection pressures given the repeated and heavy consumption of antibiotics^1^. Even though we have witnessed a significant improvement in global health, there are still millions out there who do not have adequate access to health services. Factors such as rising incomes, incessant infectious diseases, poor or marginalized populations having limited access to primary health care, and consumption of antibiotics without any prescription have been compounding the problem of antimicrobial resistance in the low-income and middle-income countries^2^. Ensuring proper sanitation, better governance, increased focus on public health care, and regulation of the private health care sector is a must to combat the spread of antimicrobial resistance^3^. While its disastrous effect on health outcomes can be understood by an estimated million deaths, the cost of treating resistant infections is estimated to reach US $100 trillion by the year 2050^4 5^. To prevent these catastrophic consequences, it is essential to develop evidence-based policies for AMR mitigation. The role of government (policymakers) is multifold in improving antibiotic stewardship policies, increasing antibiotic use surveillance, and financing these along with funding the development of new drugs ^6 7^.^8^ There arises a need to formulate governance frameworks to help the policymakers in designing, monitoring national action plans for tackling antimicrobial resistance at all levels: local, regional, national and global. It is thus essential to understand and quantify the roles of these factors in the development of resistance. Tackling antibiotic resistance is a high priority for WHO. A global action plan on antimicrobial resistance^9^, including antibiotic resistance, was endorsed at the World Health Assembly in May 2015. The global action plan aims to ensure the prevention and treatment of infectious diseases with safe and effective medicines. WHO has been leading multiple initiatives to address antimicrobial resistance like World Antimicrobial Awareness Week, The Global Antimicrobial Resistance Surveillance System (GLASS)^10^, Global Antibiotic Research and Development Partnership (GARDP)^11^, Interagency Coordination Group on Antimicrobial Resistance (IACG)^12^ with the motivation to improve awareness and understanding of antimicrobial resistance, to strengthen surveillance and research, to reduce the incidence of infection, to optimize the use of antimicrobial medicines and to ensure sustainable investment in countering antimicrobial resistance. A political declaration endorsed by Heads of State at the United Nations General Assembly in New York in September 2016 signalled the world’s commitment to taking a broad, coordinated approach to address the root causes of antimicrobial resistance across multiple sectors, especially human health, animal health, and agriculture. WHO is supporting the Member States to develop national action plans on antimicrobial resistance, based on the global action plan. Primary health care can play a vital role in tackling antimicrobial resistance. Community engagement and empowerment are essential to prevent common health problems without the unnecessary use of antimicrobials. Multisectoral action on antimicrobial resistance to limit the usage of antibiotics in the agricultural sector and ensuring equitable and good quality primary health care to all can act as an effective response to the antimicrobial resistance ^13 14^.

In this work, we quantified the relationships between a wide range of factors, from governance, healthcare access, finance, population density, to the type of sample and gender, and Global AMR. We created a compendium of Bayesian AI models to predict drug-combination patterns that are most likely to mitigate AMR in given machine learning models for resource, drug-combinations, Priority pathogen list, and Critically Important Antibiotics. Also, we used supervised machine learning models and predicted the susceptibility of antibiotics. Finally, we vetted our findings with clinicians and policymakers and created the AMR stewardship web application which can be accessed across the globe http://antibioticsteward.tavlab.iiitd.edu.in/.

## Methods

In this study, we developed a pipeline **(Figure 1)** to understand the robust connection amongst AMR, disease burden, governance, finance, and other socioeconomic indicators. We then used our results to better understand and predict the susceptibility of antibiotics amongst critical pathogens.

**Figure 1:**
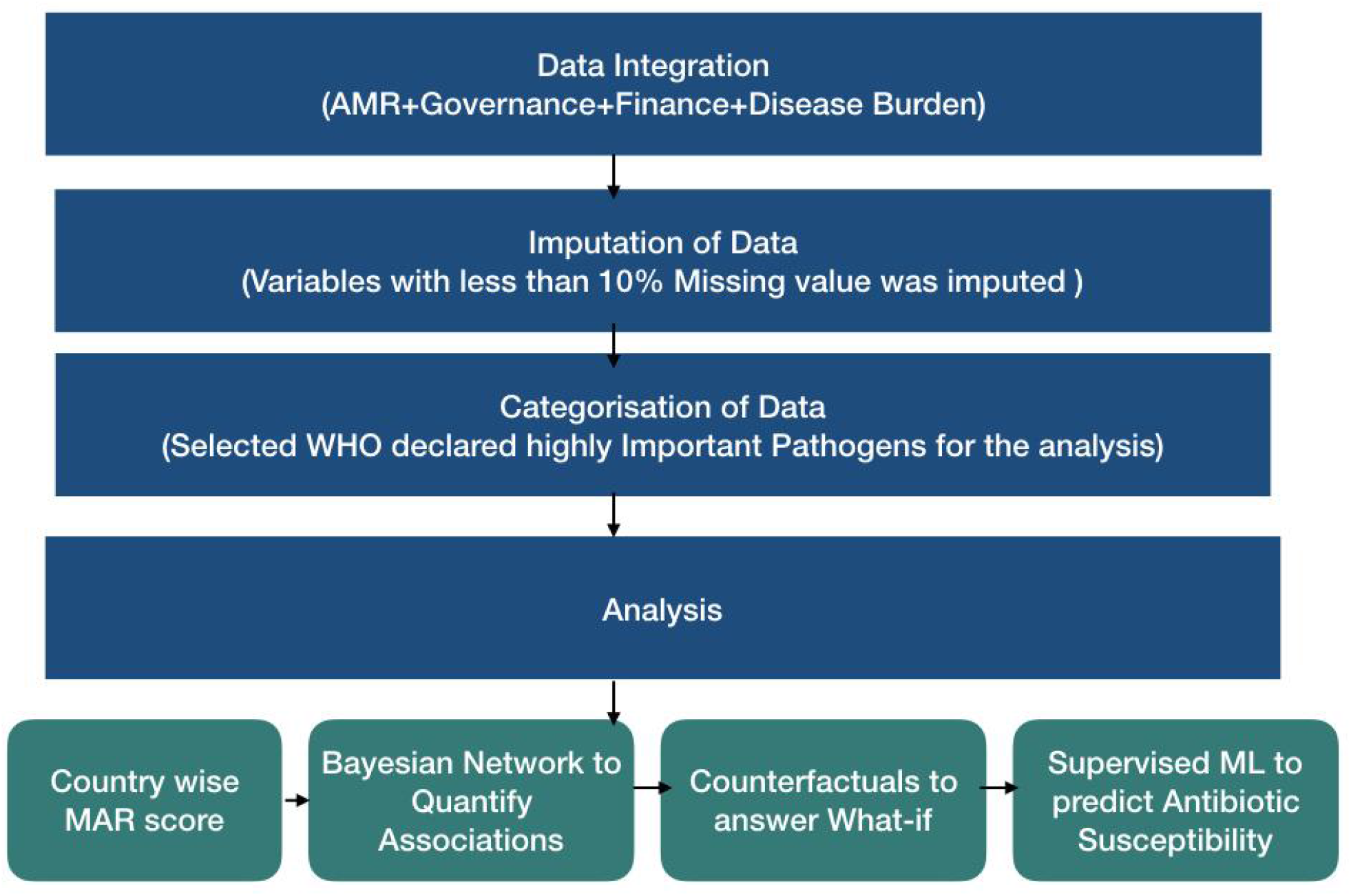
Flowchart showing pipeline used in our analysis. Firstly country level socio-economic, socio-demographic, Environmental, Food Consumption, Disease Burden and Governance data was integrated with isolated level AMR datasets. Then the Structural Model was used to identify most actionable factors of AMR, then these actionable factors were feeded to predict antibiotic susceptibility using Supervised Machine Learning Models.

### Data Integration

The AMR base data was extracted from the AMR surveillance competition ^15 16^ which had 633820 isolates. The characteristics for the same have been displayed in **(Table 1, Figure 2, Supp Fig 1)**. We then integrated the AMR data with the WGI (World Governance Indicators)^17^ data, the GBD (Global Burden of Disease Study) data^18^, and the Finance data.

**Table 1:** AMR Data was Spatio-temporal spanned from 2004 to 2017 for 70 different countries. Table represent the gender wise sample distribution for different years and also different age-groups.

|  | Female | Male | Overall |
| --- | --- | --- | --- |
| **Gender** |  |  |  |
|  | 278128 (43.88%) | 348136 (54.93%) | 633820 |
| **Age.Group** |  |  |  |
| **0 to 2 Years** | 15329 (42.83%) | 20012 (55.91%) | 35793 |
| **13 to 18 Years** | 6348 (47.06%) | 7043 (52.21%) | 13490 |
| **19 to 64 Years** | 130162 (44.04%) | 163201 (55.22%) | 295537 |
| **3 to 12 Years** | 12613 (46.88%) | 14052 (52.22%) | 26907 |
| **65 to 84 Years** | 86561 (41.63%) | 119801 (57.62%) | 207922 |
| **85 and Over** | 23406 (54.29%) | 19364 (44.91%) | 43114 |
| **Year** |  |  |  |
| **2004** | 9433 (46.93%) | 10655 (53.01%) | 20101 |
| **2005** | 10473 (48.04%) | 11274 (51.71%) | 21801 |
| **2006** | 13967 (46.65%) | 15876 (53.03%) | 29940 |
| **2007** | 18264 (45.70%) | 21409 (53.57%) | 39964 |
| **2008** | 16303 (44.33%) | 19925 (54.18%) | 36773 |
| **2009** | 18575 (44.55%) | 22514 (54.00%) | 41692 |
| **2010** | 14476 (44.59%) | 17341 (53.42%) | 32462 |
| **2011** | 11622 (44.66%) | 13931 (53.53%) | 26023 |
| **2012** | 22432 (42.16%) | 29477 (55.40%) | 53206 |
| **2013** | 29962 (42.73%) | 38850 (55.40%) | 70125 |
| **2014** | 30492 (43.23%) | 39482 (55.98%) | 70529 |
| **2015** | 27992 (43.21%) | 36104 (55.73%) | 64785 |
| **2016** | 29744 (42.74%) | 39290 (56.45%) | 69598 |
| **2017** | 24393 (42.93%) | 32008 (56.33%) | 56821 |

**Figure 2:**
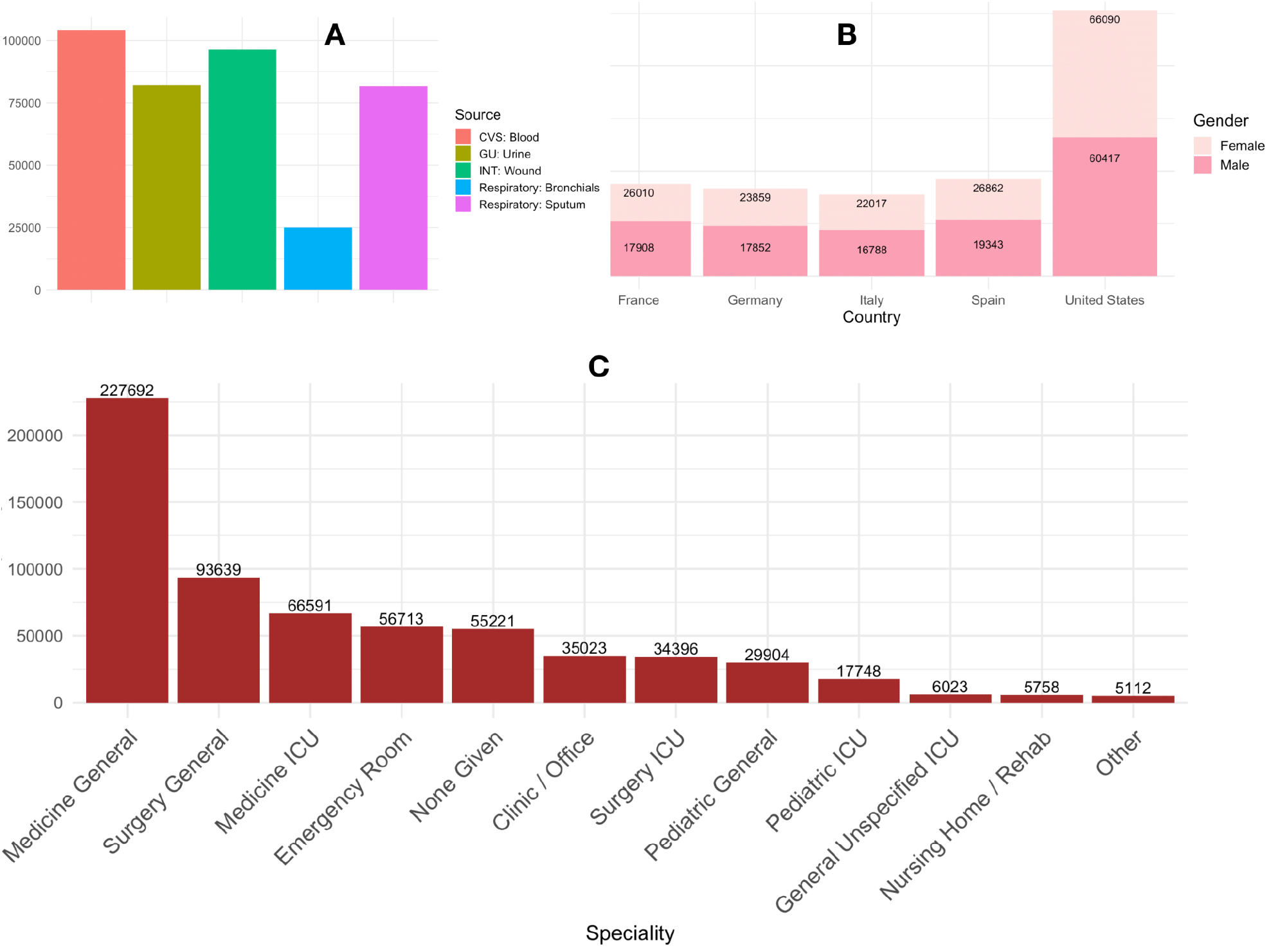
A) Bar plots showing Frequency distribution of sample collected from different sources, sample was collected from 116 different sources here we have plotted top five highest frequency sources. B) In this study data was collected from 70 middle and high income countries, here we have plotted gender wise frequency distribution of top 5 highest frequency countries. C) Bar Plot showing sample speciality of samples.

WGI data covers six dimensions of governance for over 200 countries over the period 1996-2018. The six dimensions of governance are: 1) Control of Corruption 2) Voice and Accountability 3) Political Stability and Absence of Violence/Terrorism 4) Government Effectiveness 5) Regulatory Quality6) Rule of Law, we picked estimate from each sheet corresponding to the above-mentioned dimensions of governance. Here, Estimate refers to the “Estimate of governance (ranges from approximately -2.5 (weak) to 2.5 (strong) governance performance)”. These were then merged with the AMR data set by country and year, thus giving us the resultant AMR WGI data set.

The GBD covariates data set consists of a total of 334 covariates data files for the period 1980-2017. Types of covariates used include socioeconomic, demographic, health system access, climate, and food consumption data. In the Gender, some covariates have information about gender specificity and some have information common for both. So in the latter case, we regenerated the same information for both genders and then combined the data in rows. Similarly, all covariates have three types of Age-Group 1) All Ages 2) In the form of intervals 3) Age-standardized in covariates where age groups are in the form of intervalsWe then grouped by age, gender, country and summarised by taking the mean. After having made these changes in the covariates files, these were then merged with the AMR data by country, age, and gender, taking only the value of covariates, thus giving us the resultant AMR WGI GBD data set.

We reshaped the finance data such as columns country, year and then merged with the above-mentioned AMR_WGI_GBD data set by country and year. We discarded all those variables which had more than 10% missing data, thus, giving us the resultant AMR_WGI_GBD_ FINANCE data set.

### Data Imputation and Discretization

We replaced the missingness in the AMR phenotypic data with ‘Not_Given’ and antibiotic with “Not_Tested” **(Supp Fig 5)** and the missingness in the variables of merged(amr_wgi_gbd_finance) datasets using a state-of-the-art Random Forest approach^19^ that took into account the associations between variables while inputting the data. After the imputation of data, we divided every numerical variable into three disjoint intervals i.e. Low, Medium, and High using a KNN based algorithm for the purpose of discretization^20^.

### WHO Declared Highly Important Pathogens Selection

Further, we filtered data as per the WHO declared critically important^21^ and highly important pathogens. The pathogens we took into consideration included Pseudomonas aeruginosa, Klebsiella pneumoniae, Escherichia coli, Acinetobacter baumannii, Enterobacter cloacae, Staphylococcus aureus. We considered their samples pertaining to high-income and middle-income countries (Table 2).

**Table 2:** Priority category of pathogens & samples in high income countries & middle income countries

| **Priority category of Pathogens** | **Pathogens** | **Samples in High Income Country** | **Samples in Middle Income Country** |
| --- | --- | --- | --- |
| Critical | Pseudomonas aeruginosa | 11373 | 13266 |
| Critical | Klebsiella pneumoniae | 11901 | 15437 |
| Critical | Escherichia coli | 14864 | 18125 |
| High | Staphylococcus aureus | 21456 | 27314 |
| Critical | Acinetobacter baumannii | 4468 | 5816 |
| Critical | Enterobacter cloacae | 7462 | 6860 |

### Analysis

We executed an exhaustive set of analysis which includes generative machine learning, discriminative machine learning and counterfactual (what-if) analysis.

### MAR Score

The MAR (Multiple Antibiotic Resistance)^22^ index of a single isolate is defined as a/b, where, a is the number of antibiotics to which the isolate is resistant to and b is the number of antibiotics tested.

### Bayesian Network Analysis

Bayesian networks ^23^ that learn the latent structure in complex data, represent it as compact graphical representations and allow for inferential and intervention modeling. Interpretability and explainability are the key challenges in AI-based decision models. Recent years have seen a revival of causal networks ^24^that can be learned directly and reliably from data as a quintessential approach towards achieving explainability and trust for complex problems faced by society. In this study, the Bayesian network was learned directly from complex multivariate data for explainable intervention modeling ^25 26^.

Firstly, we took all the variables of WGI GBD FINANCE AMR and learned a one-time Bayesian network. Then the Markov blankets of all antibiotics were calculated from the one-time network. The final network was learned considering only the Markov blanket variables. The inferences were then calculated from this network.

### What-if Analysis

We imagined the hypothetical situation of the variables and performed counterfactual analysis ^27^using the R package Counterfactual^28^ to answer the hypothetical ‘what if’ questions. For example, what would be the resistance of antibiotics if countries with poor health system access prevailed the characteristics of countries which have advanced health system access.

### Prediction of Antibiotic Susceptibility using Supervised Machine Learning

Identification of isolates susceptible to certain antibiotics is essential in fighting against antibiotic-resistant pathogens. So we extracted GBD, WGI, finance indicators (which were in the Markov blanket of antibiotics in the Pathogen wise Bayesian network), demographic and clinical information of isolates as a predictor of antibiotic susceptibility. Data was partitioned into training (80%) and testing (20%) sets and the class imbalance was corrected using the Synthetic Minority Oversampling Technique(SMOTE)^29,30^ Different supervised machine learning models Random-Forest(RF), Support vector machine (SVM), logistic, naive-Bayes, were learned for predicting the response to mental health indicators using the Scikit-learn library in Python.

## Results

To identify the Actionable Global determinants of AMR, we first recognised the pattern of AMR spread across the globe. We took into consideration the WHO declared critically important pathogens and performed a network analysis to identify a pathogen wise AMR mitigator. For each identified mitigator, we determined the impact in Middle and High-income countries separately. This was followed by a counterfactual analysis to measure the hypothetical inferences. Finally, using identified GBD, WGI, and finance determinants we predicted the susceptibility of antibiotics in the selected pathogens.

### Global Prevalence of Multiple Antibiotic Resistance of Critically Important Pathogens

To understand the spread pattern of Antibiotic resistance of different pathogens, we calculated the Multiple Antimicrobial Resistance (MAR) countrywise **(Figure 3)**. This revealed that every pathogens had a different pattern of resistance spread. For Enterococcus, MAR was found in the range [0.24, 0.57] with the highest value in Vietnam (0.57) and the lowest value in Venezuela (0.24). For Acinetobacter baumannii, MAR value was found in the range (0, 0.86) with the highest value in Vietnam (0.86). For E coli, MAR value was found in the range [0.032, 0.45] with highest values in Indonesia (0.45), India (0.40) and the lowest value in Norway (0.032). For Klebsiella pneumoniae, MAR was found in the range of [0.09, 0.64] with the highest value in Serbia (0.64) and the lowest value in Japan (0.09). For Pseudomonas aeruginosa, MAR value was observed in the range [0.016, 0.75] with the highest value in El Salvador (0.75) and the lowest value in Norway (0.016).

**Figure 3:**
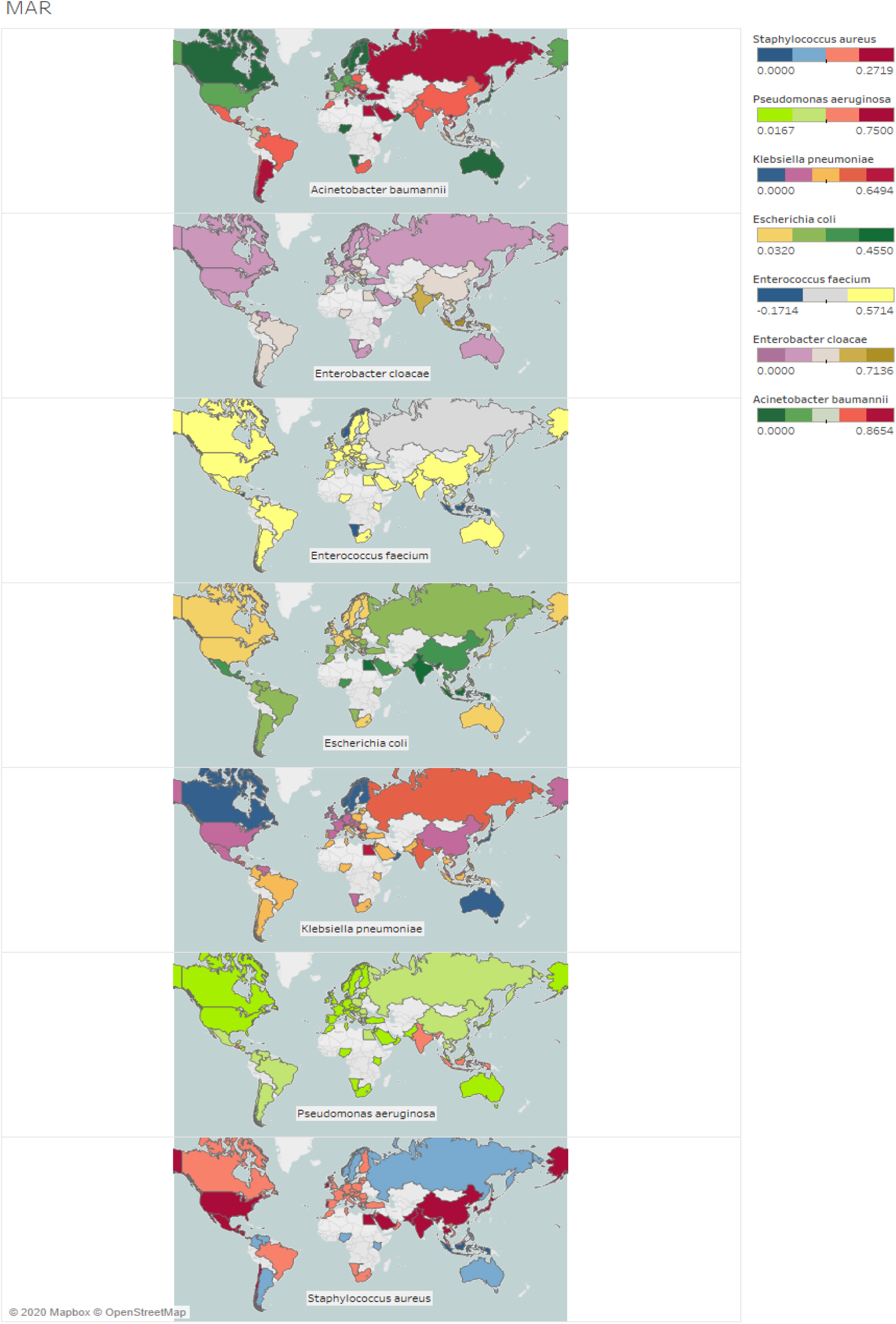
Global prevalence of antibiotic resistance in different pathogens. We calculated the MAR score for the isolated then calculated the country-wise mean of MAR score, which explained the resistance pattern.

### Component Level Bayesian Network Revealed Strong Connection of Antibiotics with Independent WGI, Finance, GBD datasets

In order to check the directional relationship among the independent datasets i.e. WGI, GBD, Finance and AMR, we performed Principal component analysis (PCA) in the Numerical data set and multiple correspondence analysis (MCA) in Categorical data sets. We picked 2 components which captured higher variance of datasets and with the help of these 2 components, on a one time network **(Figure 4)** we found a strong association of governance, finance and GBD principal components with Antibiotics components.

**Figure 4:**
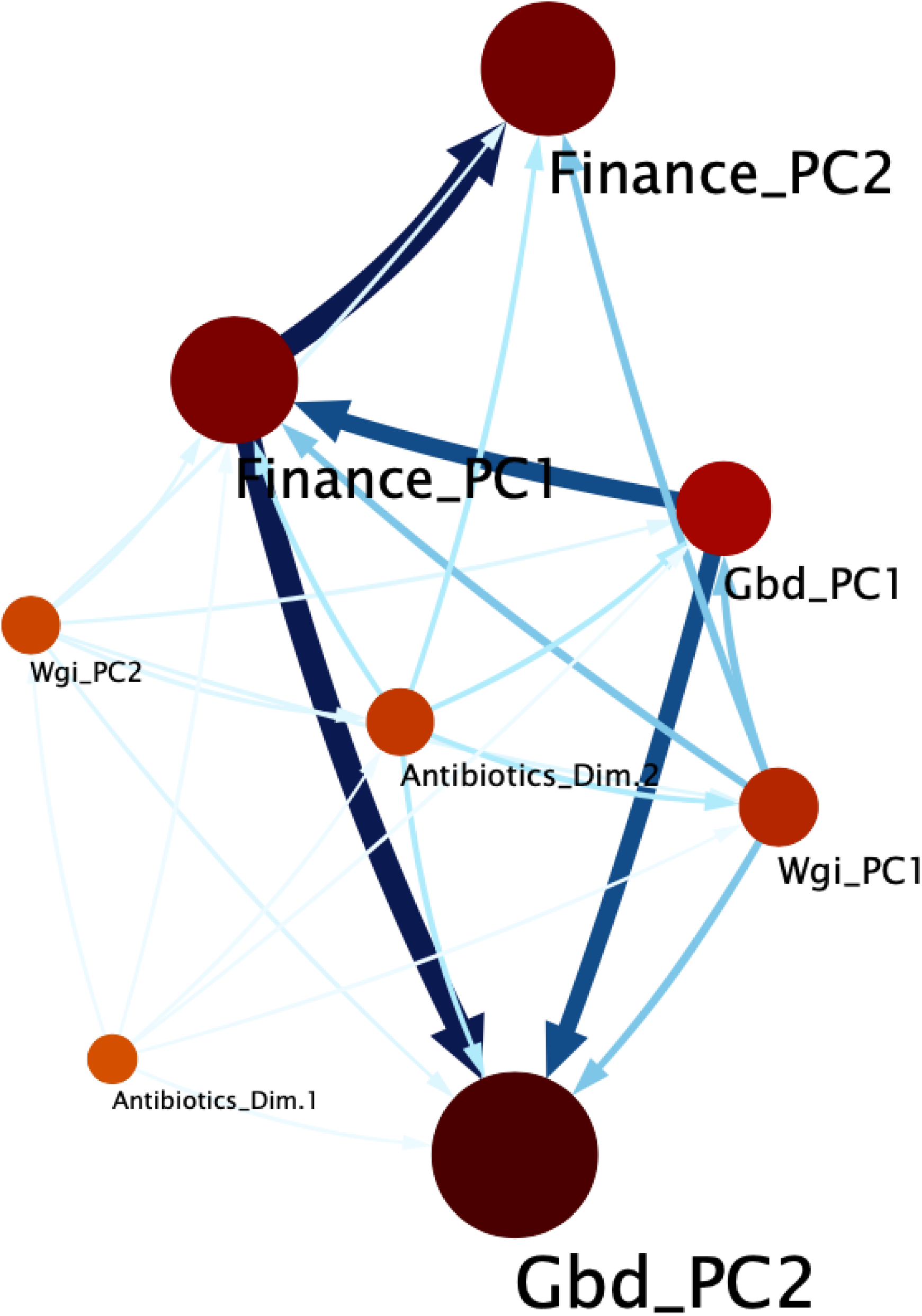
Bayesian network of principal components reveals strong connection of AMR with WGI, Finance, GBD.

### Probabilistic Impact Quantification of Governance, Finance, Socioeconomic and Burden of Disease on Antibiotic Resistance

For WHO declared critical pathogens we learned a robust bootstrapped network. From the Network Inference governance of the country and health system access, we found a strong influence of AMR in all pathogens. We found the probability of cefepime resistance in E coli pathogens to be 21% higher in a country where voice and accountability **(Refer Supplementary Definitions)** was low as compared to a country where voice and accountability was high. The probability of piperacillin resistance in Acinetobacter baumannii pathogens is 50% lower in a country where there is a high level of government effectiveness. The probability of cefepime resistance is 12.98% lower in the pathogens Pseudomonas aeruginosa and 13% lower in the pathogens Staphylococcus aureus in a country where there is high level of government effectiveness as compared to a country having a low level of government effectiveness. Similarly, the probability of amikacin resistance was 12.17% lower in the pathogens Pseudomonas aeruginosa and 11.6% lower in the pathogens Staphylococcus aureus in a country where there was a high level of government effectiveness as compared to a country having a low level of government effectiveness. We also found that good health system access in a country significantly decreases ceftriaxone and meropenem resistance in many pathogens of bacteria. The probability of ceftriaxone resistance in countries with good health system access was observed to be 14.74% lower in E Coli, 36% lower in Acinetobacter baumannii, 12.47% lower in Enterobacter Cloacae, 15% lower in Klebsiella pneumoniae while the probability of meropenem resistance in Klebsiella pneumoniae was observed to be 3.43% higher in countries with good health system access. Improvements in water sanitation and hygiene (WASH) are critical elements of preventing infections and reducing the spread of antimicrobial resistance (AMR) as identified in the Global Action Plan to combat AMR. In our analysis, we found out that the probability of cefepime resistance in Klebsiella pneumonia was 19.87% higher in countries where the level of Unsafe Wash Sanitation was high. Our analysis also quantified the effect of fruit consumption on cefepime resistance in Klebsiella pneumoniae. High food consumption decreased the chance of cefepime resistance by 13.04% **(Table 3)**.

**Table 3:**
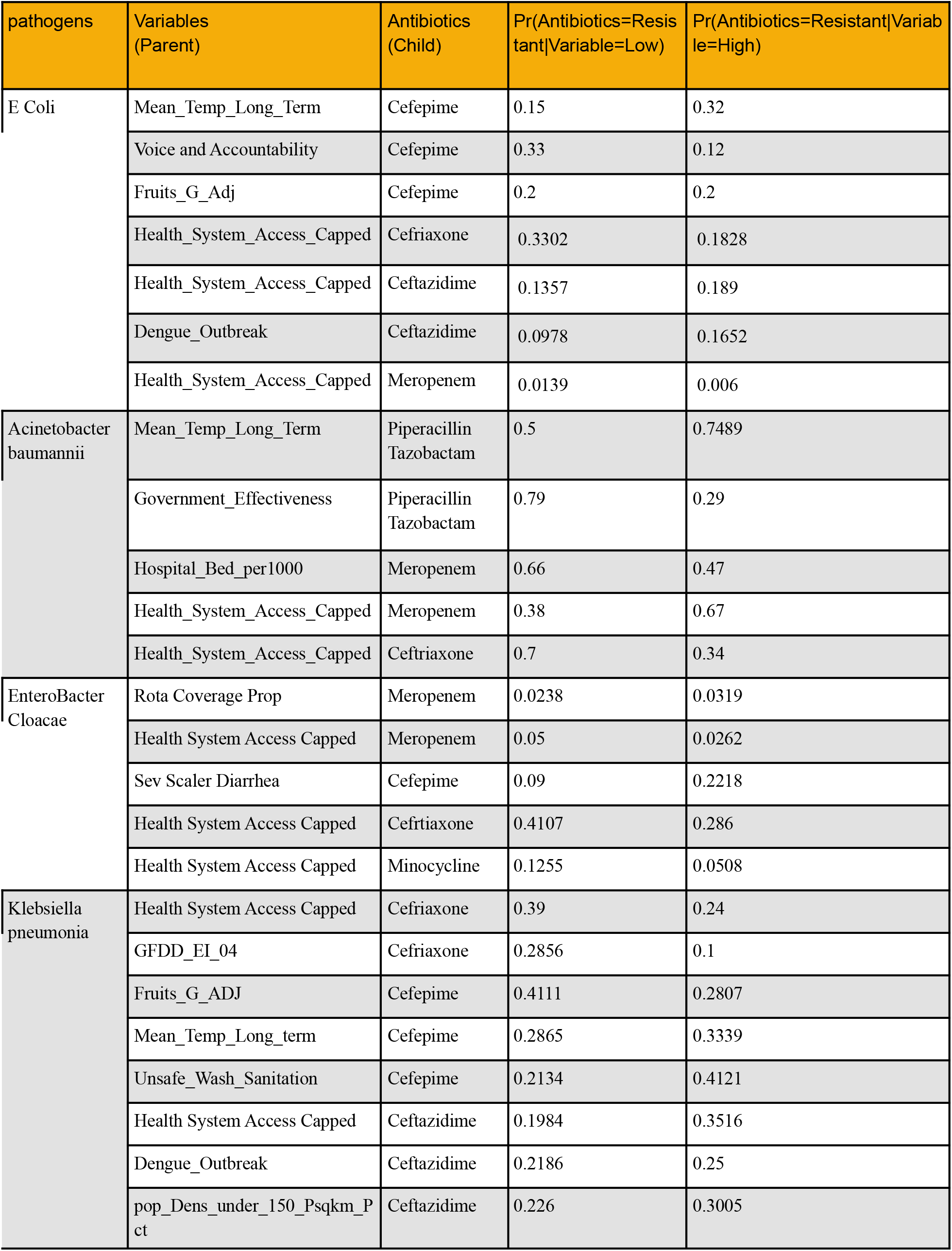
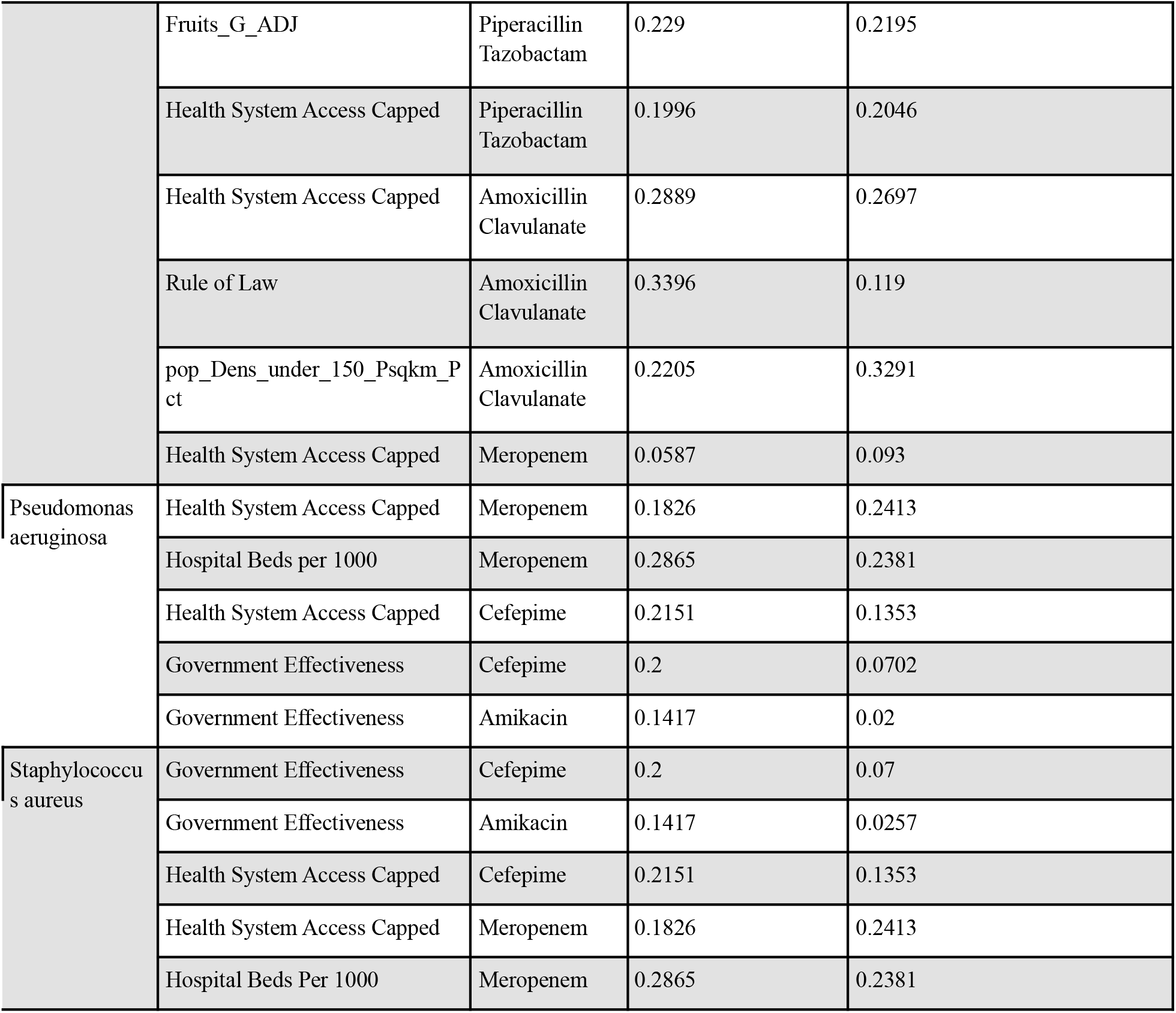
Inferences from pathogen wise bayesian network.

### Counterfactual Analysis Quantified the Reduction in Ceftriaxone Resistance if Countries with Poor Health System Access follow Advanced Health system Access

From Bayesian network analysis, we observed that the health system access has impacted the Ceftriaxone resistance with high magnitude consistently for all pathogens So, we performed a counterfactual analysis on the ceftriaxone resistance in the Middle and High-income countries. For the pathogen Enterobacter Cloacae, counterfactual effect (what would be the ceftriaxone resistance if poor health system follows the characteristics of advanced health system access) of health system access on ceftriaxone e resistance was observe **(Figure 5)** to be consistent in the Middle and High-income countries with median quantile effect in Middle-income countries to be 0.007 with 95% CI [-0.05, 0.0.7] and in High-income countries to be 0.02 with 95% CI [0.003, 0.04]. For Acinetobacter baumannii and E Coli, we found the counterfactual effect of health system access on ceftriaxone in the Middle-income countries to be much higher than that in the High-income countries. The median quantile effect in the Middle-income countries was observed to be -0.9 with 95% CI [-0.17,-0.02] in the case of Acinetobacter baumannii and -0.03 with 95%CI [-0.16, 0.10] in the case of E Coli. The median quantile effect in the High-income countries was observed to be 0.05 with 95% CI [-0.03, 013] in the case of Acinetobacter baumannii and -0.004 with 95% CI [-0.03, 0.02] in the case of E Coli. although we have performed same counterfactual analysis for meropenem resistance which was also found be highly associated with health system access however we have not found any monotonic trend in this case **(Supp Fig 4)**

**Figure 5:**
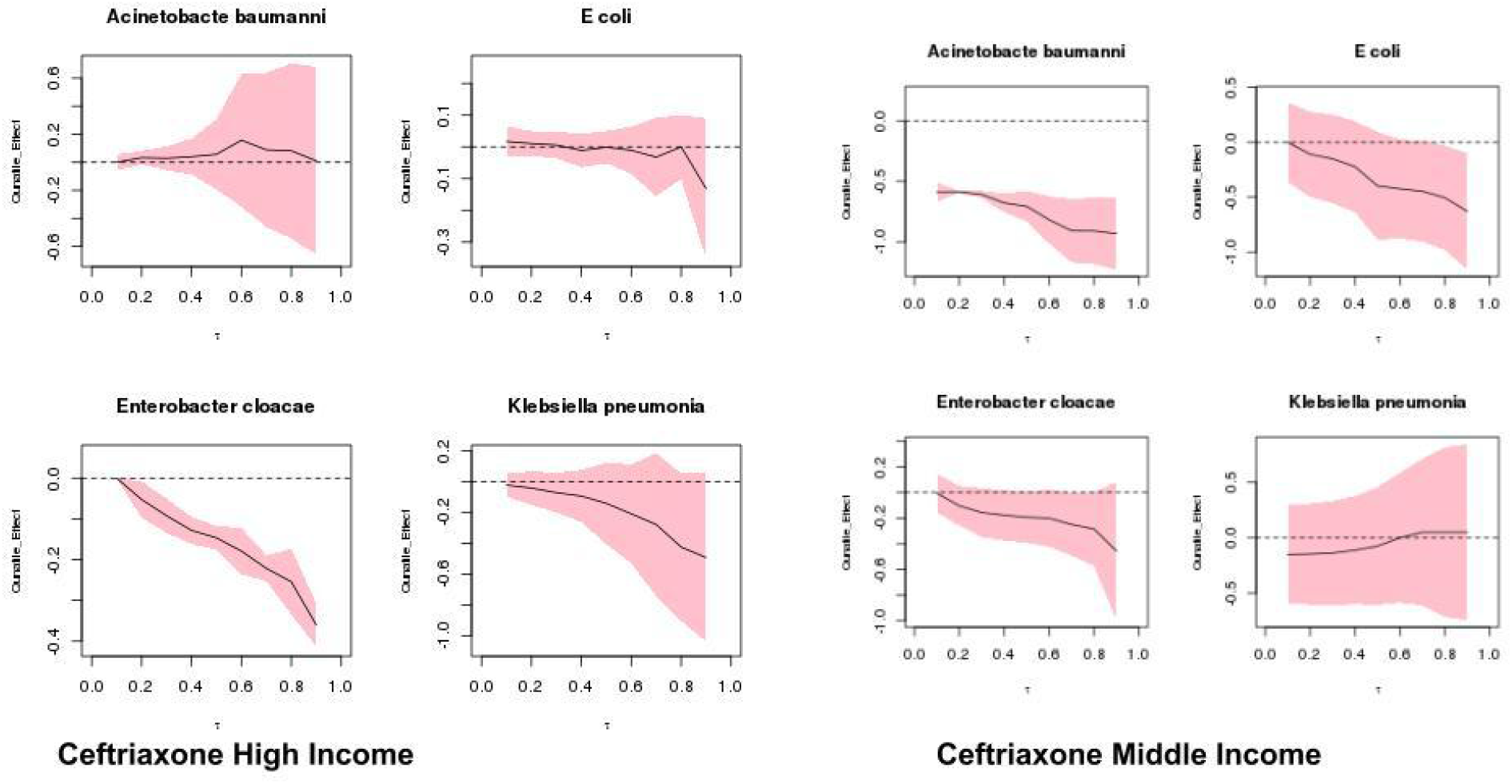
Counterfactual analysis answered the question what could be resistance of Ceftriaxone and if countries with “Poor health system access” have “High health system access”. Analysis was performed separately for the middle and high income countries.

### Prediction of Antibiotic Susceptibility

Overall we found that the Random Forest Model was the best performing model **(Supp Fig 2)**. Amongst all the pathogens, interestingly our prediction models have performed best for the Staphylococcus aureus **(Figure 6)** which is known as the most dangerous amongst the common staphylococcal bacteria that often cause skin infections. For Staphylococcus aureus our model predicted ceftaroline and oxacillin with highest AUROC 0.94 and 0.89 respectively (from RF model). We also found that our model performed well for the cefepime and ceftazidime susceptibility prediction for Klebsiella pneumoniae with AUCROC 0.88, 0.92 respectively (from RF model). From our prediction models we have also found that meropenem susceptibility is highly predictable for the Escherichia coli (AUROC=0.93) compared to other pathogens like Pseudomonas aeruginosa (AUROC=0.75), Klebsiella pneumoniae (AUROC=0.81), Acinetobacter baumannii (AUROC=0.75). On further analysis of the feature importance of prediction model (RF) for meropenem susceptibility we found that Echerichia coli and, interestingly, health system access of a country to be the most important feature **(Supp Fig 3)**. Although our model has not shown high performance for pathogens like Pseudomonas aeruginosa, overall our models have shown decent and actionable performance.

**Figure 6:**
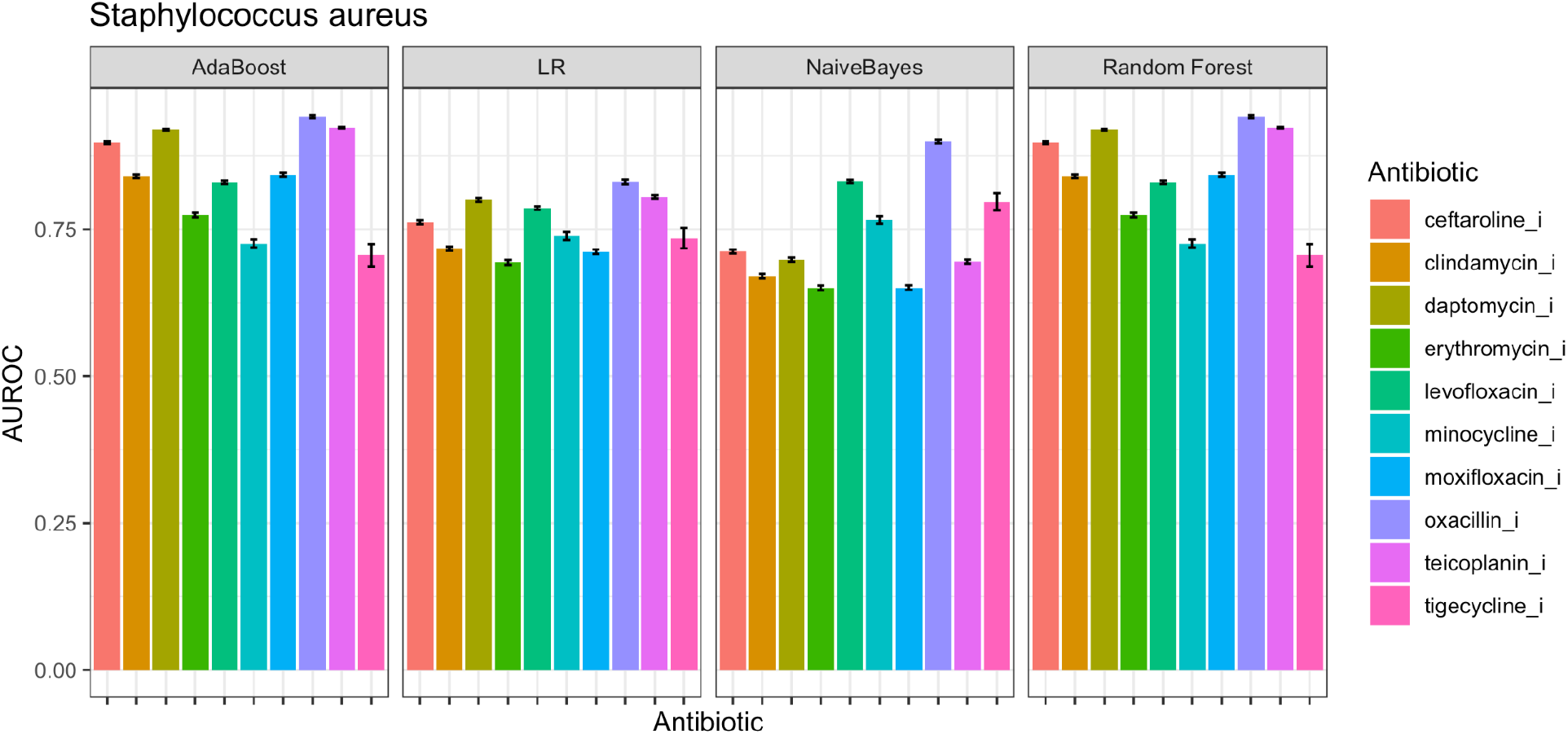
AUROC of prediction models for Staphylococcus aureus. Error bar in the figure denotes the 95% CI AUROC.

## Discussion

Antibiotic resistance is rising to alarmingly high levels in all parts of the world. New resistance mechanisms are emerging and spreading globally, threatening our ability to treat common infectious diseases^31^. A growing list of infections such as pneumonia, tuberculosis, blood poisoning, gonorrhoea and foodborne diseases are becoming harder and sometimes impossible to treat as antibiotics become less effective^32^. The emergence and spread of the resistance are worse in those regions where the antimicrobial medicines can be bought for human or animal use without a prescription. Similarly, in countries without standard treatment guidelines, antibiotics are often over-prescribed by health workers and veterinarians and overused and misused by the public. Certain economic and governance factors such as Political Stability and Absence of Violence/Terrorism, Control of Corruption, Voice and Accountability, Government Effectiveness, Regulatory Quality and Rule of Law play a key role in designing such frameworks which can help curtail the spread of Anti-microbial resistance.

By consolidating four different datasets (the AMR, GBD, WGI and Finance data sets) into one, this study aims to find the actionable global determinants. Taking into consideration the prevalence of MAR score across the globe, our findings report that MAR score is non-uniform across the globe for critically important pathogens. This finding calls upon devising policies that are effective at each country level.

Using AI techniques like Bayesian Network Analysis, we found a strong connection of AMR with the WGI, Finance and GBD datasets which motivated us to proceed with Counterfactual analysis of Ceftriaxone. As per the results, the counterfactual effect of Health system Access on Ceftriaxone in the Middle-income countries turned out to be much higher than that in the High-income countries^33^.

With the on-going advancements in medicine, we have breakthrough treatment techniques from complex organ transplants to robotic surgeries. All of these have been possible by keeping bacterial infections under control. But the rising instances of Antibiotic resistance, which are accelerated by the misuse and overuse of antibiotics, may render simple bacterial diseases untreatable. The time calls upon strategies at every stratum of the society to reduce the impact and limit the spread of the resistance. Global governance systems^34^ and finance regulators, industrial stakeholders, medical experts and scientists need to stand united to tackle this problem first hand. The Global Reference List ^35^includes priority indicators pertaining to four domains namely health status, service coverage, health systems and risk factors which countries can use to monitor their health priorities at national and sub-national levels. Providing timely access to the healthcare system, judicious and lawful uses of medicinal resources, awareness regarding public hygiene with strict action against unlawful practices, such as drug distribution without prescription, should be taken at all costs.

There are a few limitations of our work. Some confounders are still missing such as government effectiveness and health-system influenced AMR. Thus, a complete pathway has not been explained due to the dearth of data. We had data only for 70 countries which included only Middle-income and High-income countries. But Governance finance-related Intervention might also be required in case of Low-income countries. Our work is focused on high-level policy-making, so it does not include biological aspects such as Microbiome role for AMR.

## Data Availability

Culture Sensitivity Data were obtained on registration to the Wellcome AMR Surveillance Prize. Analysis and results used in this publication were generated as part of the Wellcome Trust Data Re-use Prize: AMR Surveillance, obtained through Synapse ID syn17009517 and with support from the Open Data Institute, Pfizer, and Sage Bionetworks. Data for Worldwide Governance Indicators, Global Burden of Disease, and Global Financial Development Database are publicly available.

https://www.worldbank.org/en/publication/gfdr/data/global-financial-development-database

https://datacatalog.worldbank.org/dataset/worldwide-governance-indicators

http://www.healthdata.org/gbd/2019

## Acknowledgments

This work was partially supported by the Wellcome Trust/DBT India Alliance Fellowship IA/CPHE/14/1/501504 awarded to Tavpritesh Sethi and the Center for Artificial Intelligence at IIIT-Delhi. Authors also acknowledge Prof. Rakesh Lodha from All India Institute of Medical Sciences, New Delhi, for his valuable inputs. We also acknowledge Mehrab Singh Gill, Arshita Bhatt. For their support in figure preparation and helping in manuscript writing.

## Notes

### Competing Interest Statement

The authors have declared no competing interest.

### Author Declarations

The ATLAS dataset (available for download at https://amr.theodi.org/programmes/atlas) is an open-access dataset on human AMR surveillance data generated by the commercial pharmaceutical company Pfizer that contains granular antibiotic susceptibility data made publicly available as per the DAVOS Declaration and United Nations Global Assembly Declaration in compliance with the IRB policies and detailed at https://amr.theodi.org/

### Summary of Updates

Language, figures, and tables improved.

