## Supplementary Material for "Capturing Global, Predicting Local for Controlling Antimicrobial Resistance: a retrospective multivariable analysis"

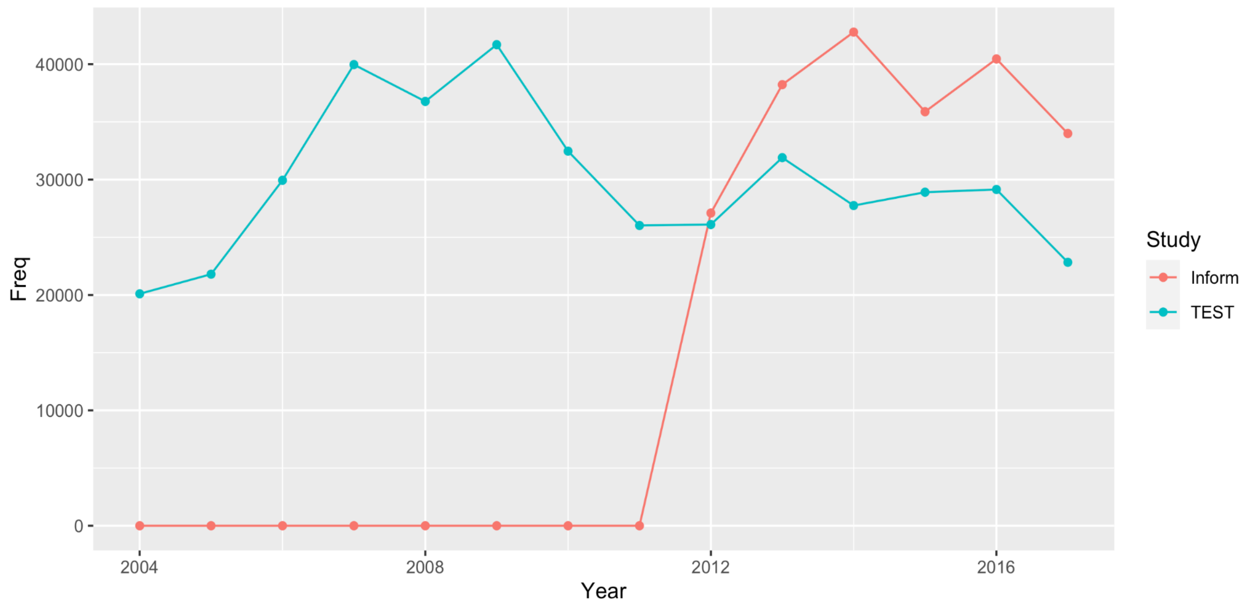


***Supp Fig 1: Data from 2 studies 1)Test (Tigecycline Evaluation Surveillance Trial) and 2) Inform (International Network for Optimal Resistance Monitoring) was used in the AMR data sets. Here we have plotted the year-wise sample size from these two studies.***

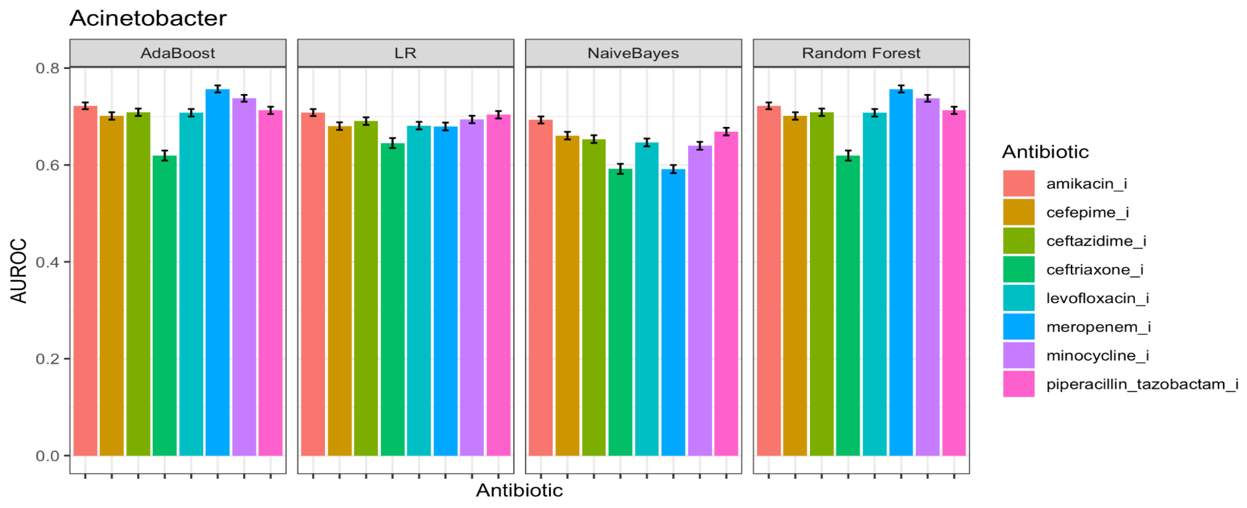


***Supp Fig 2.1: AUROC for the antibiotic susceptibility prediction for the Acinetobacter.***

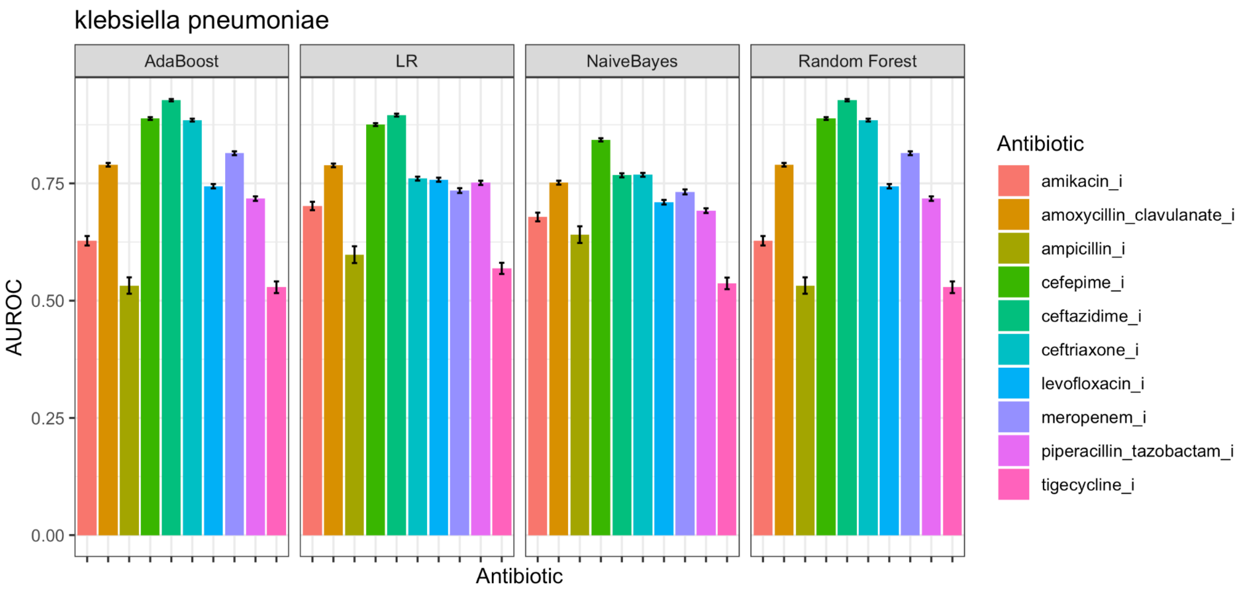


***Supp Fig 2.2: AUROC for the antibiotic susceptibility prediction for the Klebsiella pneumoniae***

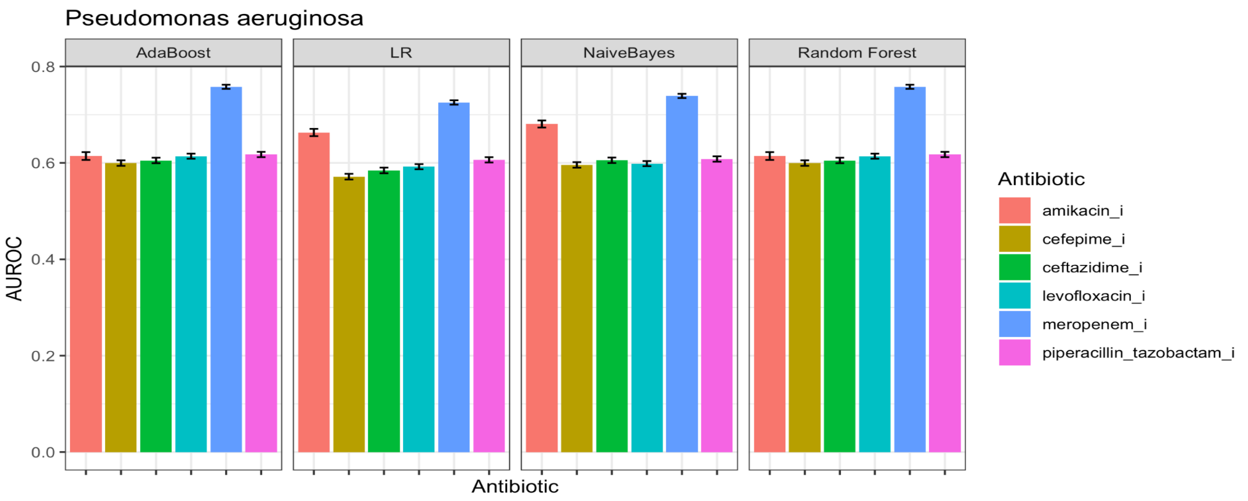


***Supp Fig 2.3: AUROC for the antibiotic susceptibility prediction for the Pseudomonas aeruginosa***

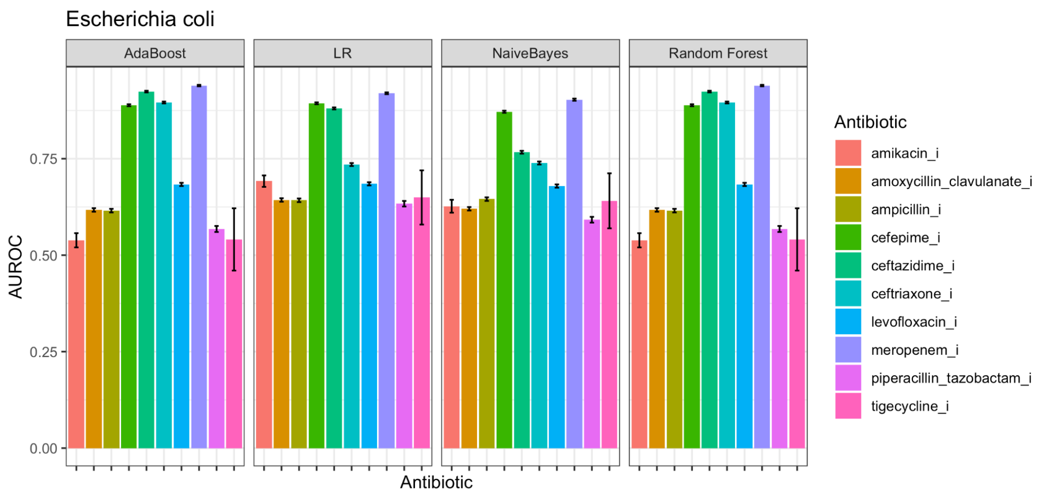


***Supp Fig 2.4 AUROC for the antibiotic susceptibility prediction for the E.coli***

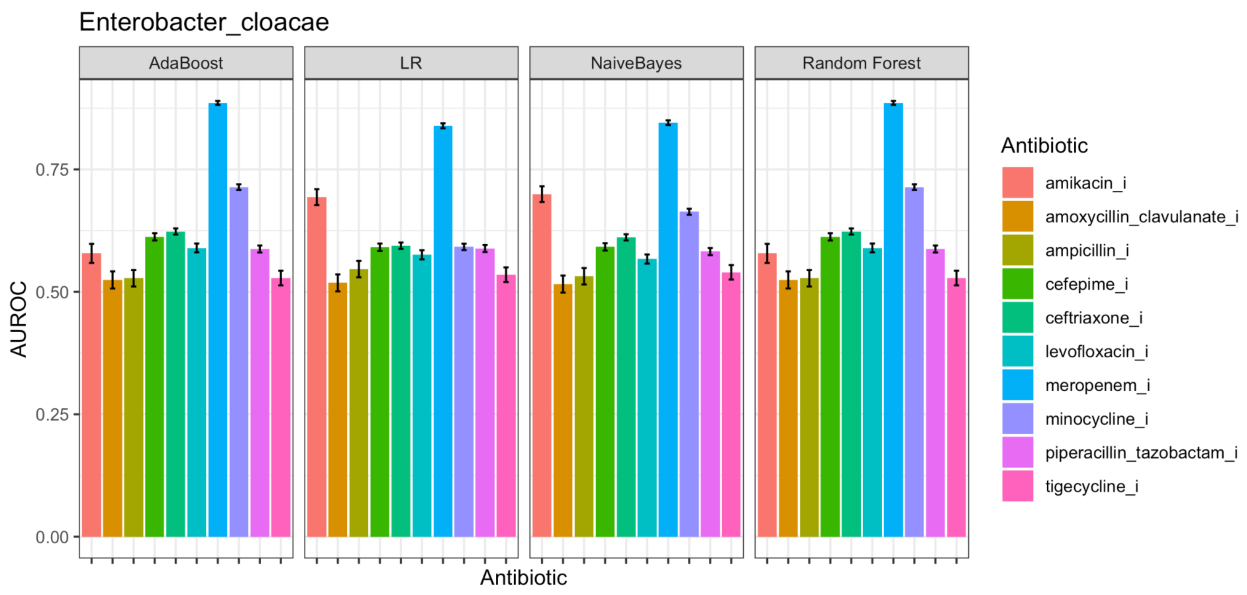


***Supp Fig 2.5: AUROC for the antibiotic susceptibility prediction for the Enterobacter cloacae***

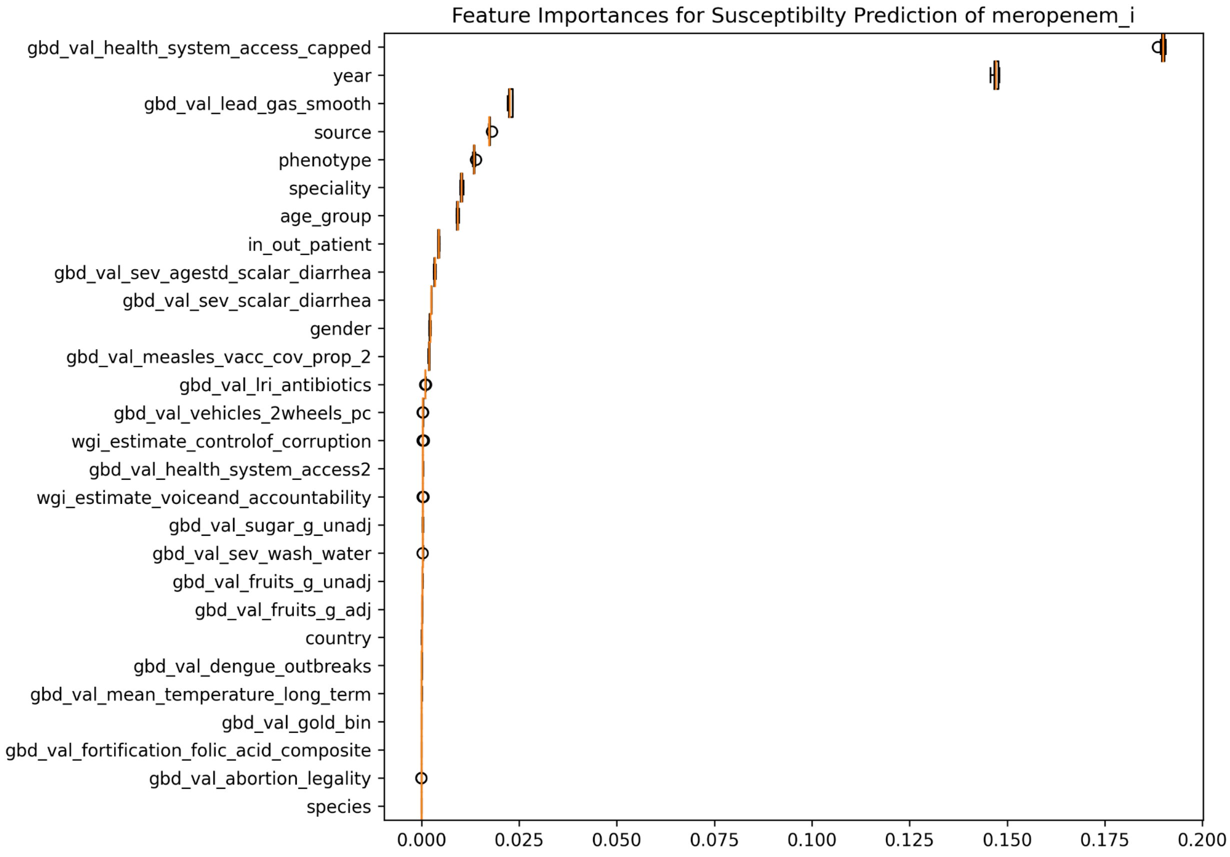


***Supp Fig 3: Feature Importance for the Prediction of Meropenem Susceptibility against E.Coli obtained from best performing the Random Forest.***

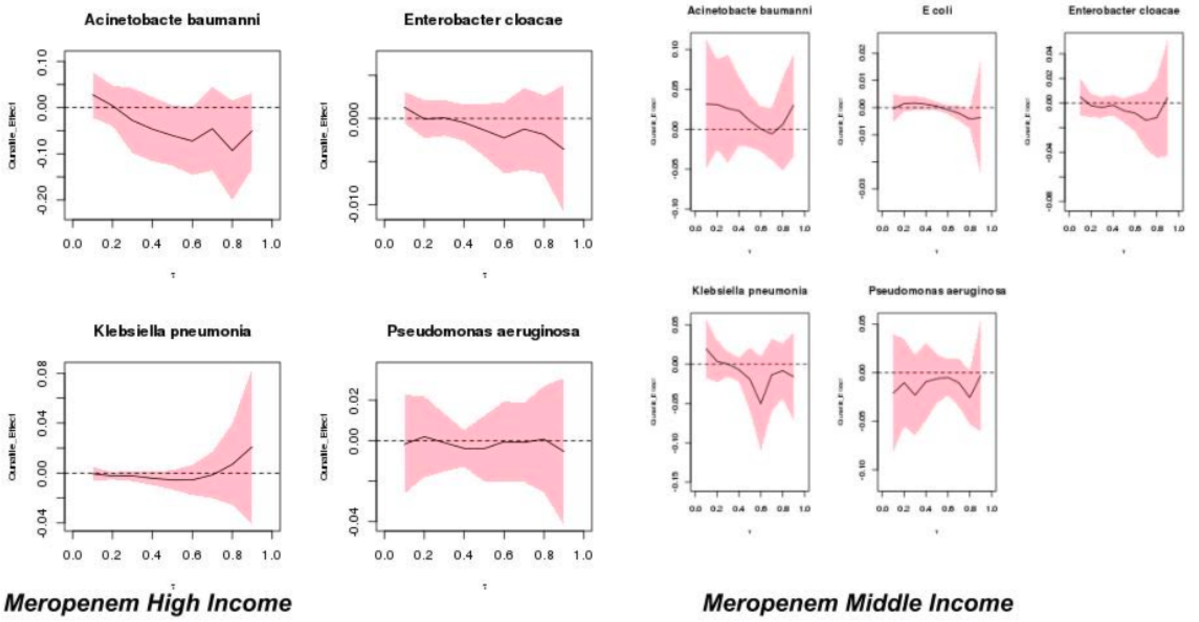


*Supp Fig 4: Counterfactual Analysis on Meropenem Resistance.*

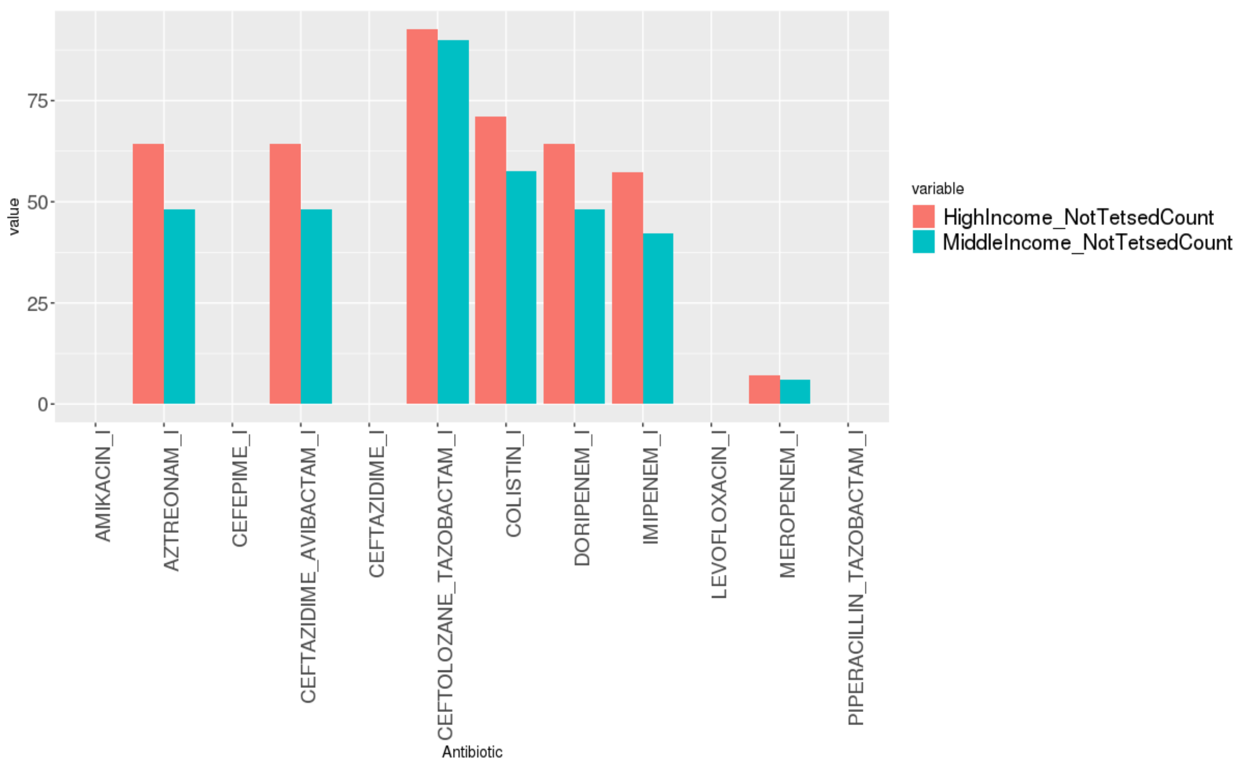


***Supp Fig 5.1: Pseudomonas aeruginosa (% of Not_Tested in Antibiotics)***

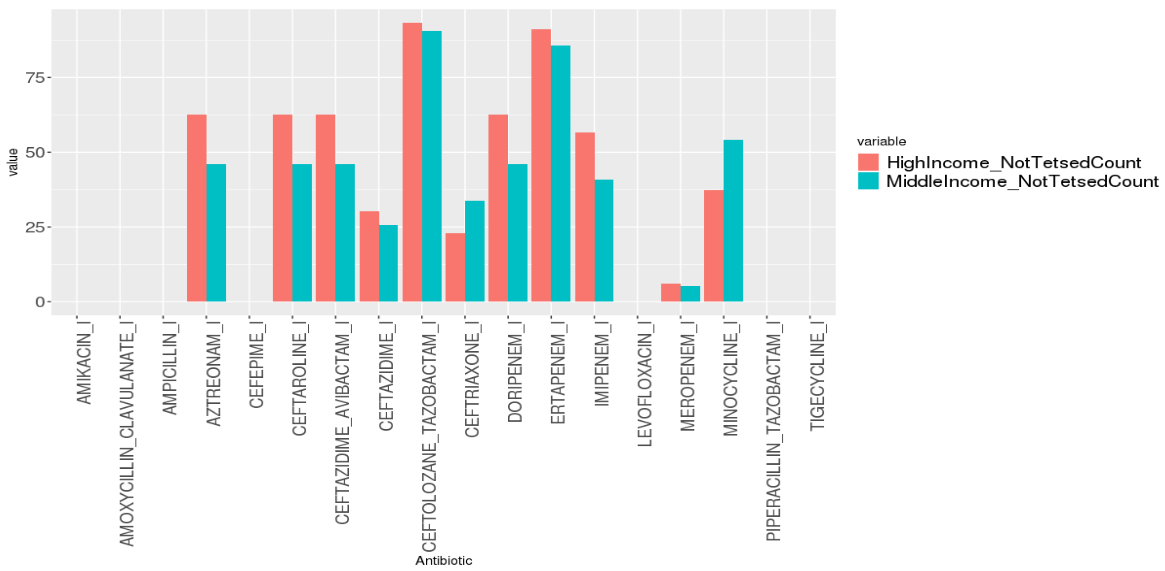


***Supp Fig 5.2: Klebsiella pneumonia: (% of Not_Tested in Antibiotics)***

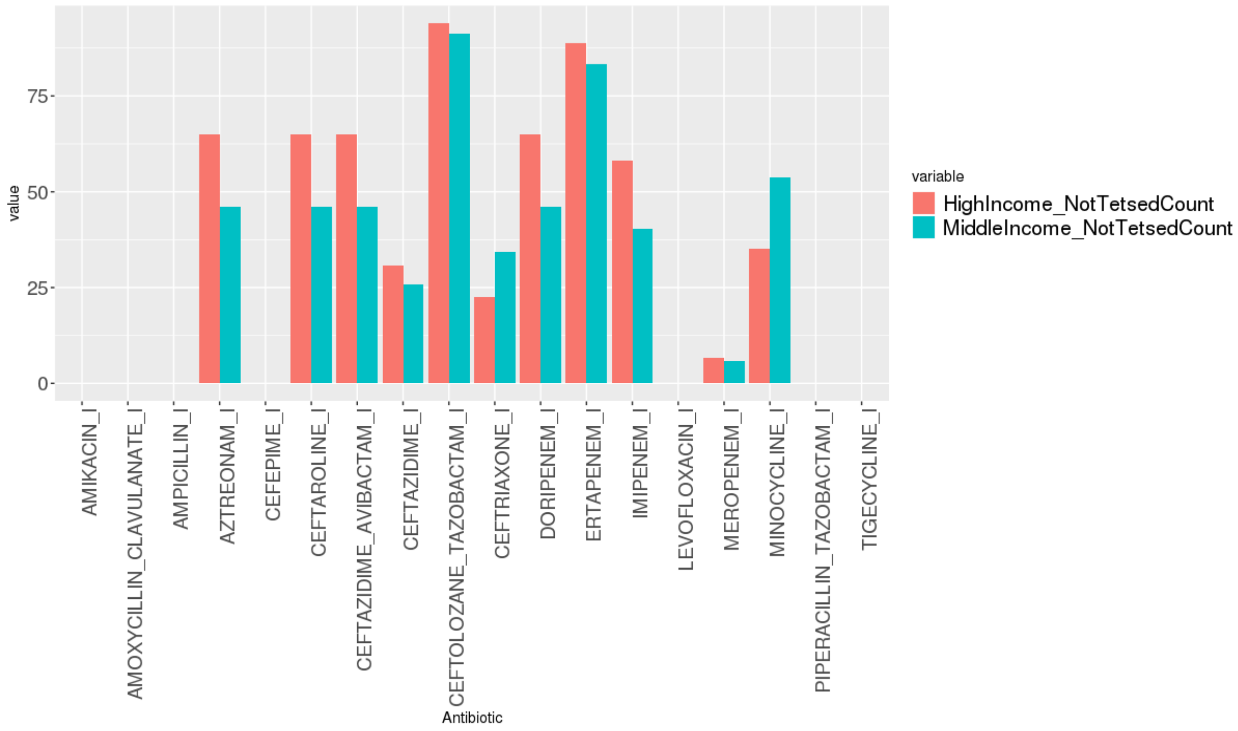


***Supp Fig 5.3:Escherichia coli (% of Not_Tested in Antibiotics)***

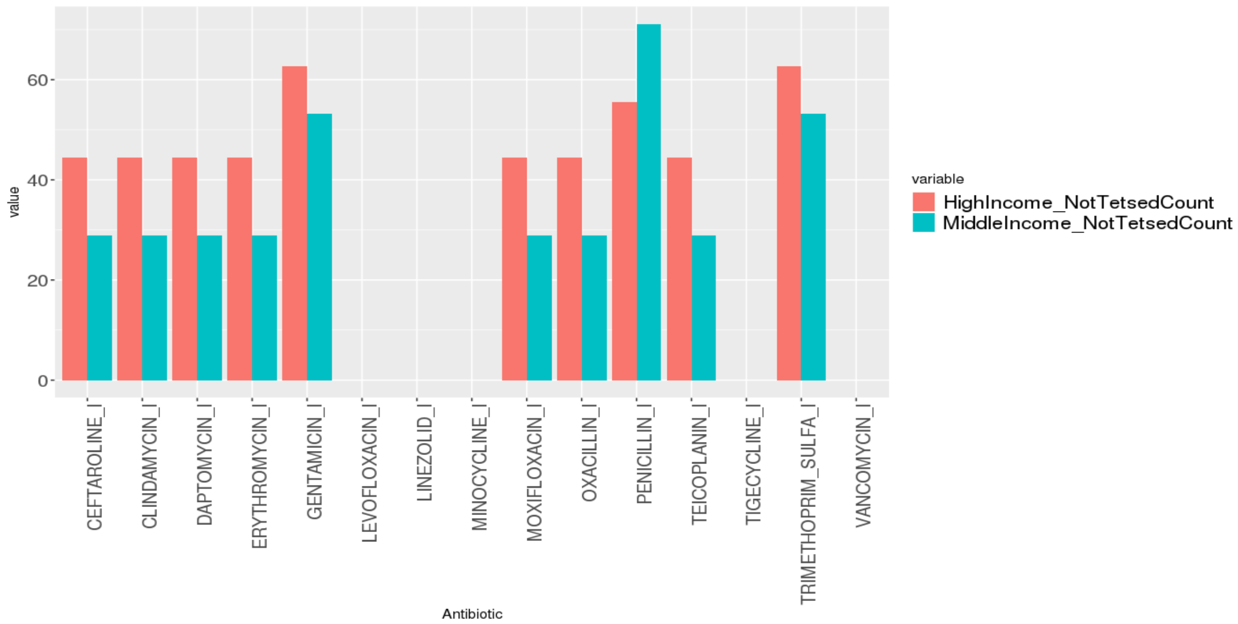


***Supp Fig 5.4:Staphylococcus aureus (% of Not_Tested in Antibiotics)***

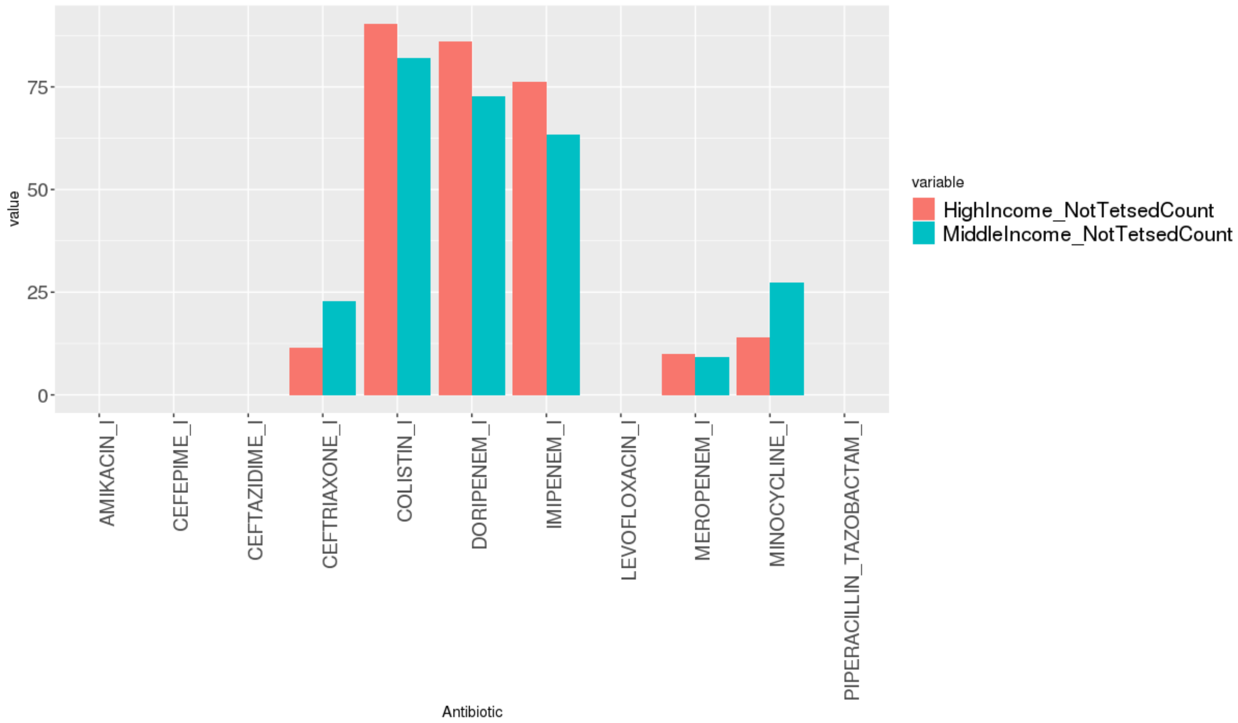


***Supp Fig 5.5:Acinetobacter baumannii (% of Not_Tested in Antibiotics)***

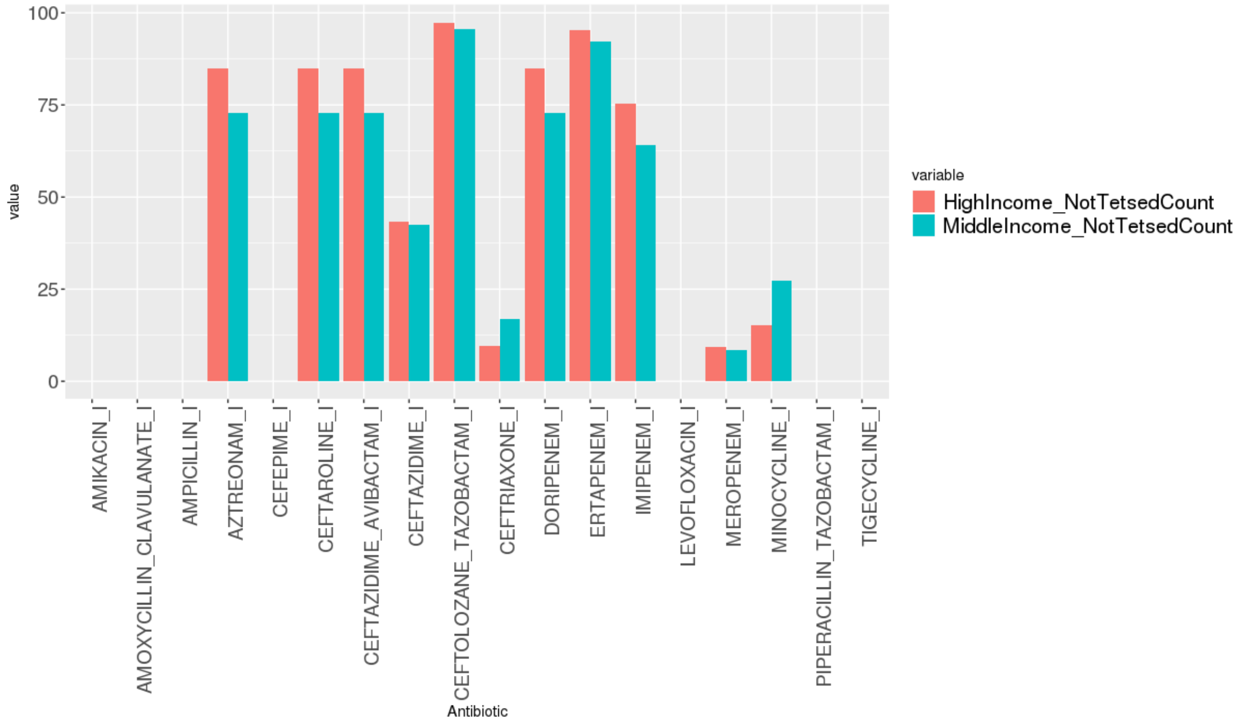

*Supp Fig 5.6:Enterobacter cloacae(% of Not_Tested in Antibiotics)*

### Supplementary Definitions :

1) **Health System Access:** A measure of health system access estimated using a principal component analysis of antenatal clinics, DTP3 immunization, measles immunization, hospital beds, in-facility delivery, and skilled birth attendance.

2) **Mean Temp Long Term:** Population-weighted mean temperature

3) **Fruits_G_ADJ**: Fruits g/p/d avail adjusted

4) **Dengue Outbreak**: A binary indicator for each country and year of dengue outbreaks; 0 = no outbreak, 1= outbreak

5) **Sev Scaler Diarrhea :** All Risk Factors SEV scalar for cause Diarrhea

6) **Rule of Law**: Rule of law captures perceptions of the extent to which agents have confidence in and abide by the rules of society, and in particular the quality of contract enforcement, property rights, the police, and the courts, as well as the likelihood of crime and violence. This table lists the individual variables from each data source used to construct this measure in the Worldwide Governance Indicators.

7) **Government_Effectiveness :** Government effectiveness captures perceptions of the quality of public services, the quality of the civil service and the degree of its independence from political pressures, the quality of policy formulation and implementation, and the credibility of the government's commitment to such policies. This table lists the individual variables from each data source used to construct this measure in the Worldwide Governance Indicators

8) **Voice and Accountability:** Voice and accountability captures perceptions of the extent to which a country's citizens are able to participate in selecting their government, as well as freedom of expression, freedom of association, and a free media. This table lists the individual variables from each data source used to construct this measure in the Worldwide Governance Indicators

9) **GFDD_EI_04:** Bank's Overhead Costs to Total Assets.
